# DETECT: Feature extraction method for disease trajectory modeling

**DOI:** 10.1101/2022.11.06.22281817

**Authors:** Pankhuri Singhal, Lindsay Guare, Colleen Morse, Marta Byrska-Bishop, Marie A. Guerraty, Dokyoon Kim, Marylyn D. Ritchie, Anurag Verma

**Affiliations:** Department of Genetics, University of Pennsylvania, Philadelphia, PA; Department of Medicine, University of Pennsylvania, Philadelphia, PA; New York Genome Center, New York, NY; Department of Biostatistics, Epidemiology, and Informatics, Perelman School of Medicine, University of Pennsylvania, Philadelphia, PA; Institute of Biomedical Informatics, University of Pennsylvania, Philadelphia, PA; Center for Precision Medicine, Perelman School of Medicine, University of Pennsylvania, Philadelphia, PA; Corporal Michael J. Crescenz VA Medical Center, Philadelphia, PA

## Abstract

Modeling with longitudinal electronic health record (EHR) data proves challenging given the high dimensionality, redundancy, and noise captured in EHR. In order to improve precision medicine strategies and identify predictors of disease risk in advance, evaluating meaningful patient disease trajectories is essential. In this study, we develop the algorithm **D**iseas**E T**rajectory f**E**ature extra**CT**ion (**DETECT)** for feature extraction and trajectory generation in high-throughput temporal EHR data. This algorithm can 1) simulate longitudinal individual-level EHR data, specified to user parameters of scale, complexity, and noise and 2) use a convergent relative risk framework to test intermediate codes occurring between a specified index code(s) and outcome code(s) to determine if they are predictive features of the outcome. We benchmarked our method on simulated data and generated real-world disease trajectories using DETECT in a cohort of 145,575 individuals diagnosed with hypertension in Penn Medicine EHR for severe cardiometabolic outcomes.

## Introduction

Recent studies have shown that patients sharing the same initial disease diagnosis can experience vastly different clinical outcomes over time (García-Olmos et al., 2012). Numerous studies have shown the potential of leveraging longitudinal data in disease risk modeling. Identifying which clinical events in an individual’s disease trajectory predict an outcome remains challenging. First, this is due to mounting EHR data, the growing scale of which makes it difficult to temporally aggregate different clinical variables in an interpretable manner. Second, patient disease trajectories are unique; a person’s health is attributable to a complex web of correlated health, genetic, and lifestyle variables. Extracting patterns of disease progression in EHR data for predictive modeling requires preserving individual-level clinical information to make population-level extrapolations. There is no gold standard approach for producing individual patient trajectories based on signals of disease pathology and progression rather than noisy data. Noise in an EHR could be driven by features such as drug side effects, external injuries, or short-term conditions driven by environmental changes - at different times, these are likely unrelated to chronic disease progression. Modeling disease trajectories for complex outcomes such as renal failure, stroke, congestive heart failure, or myocardial infarction is essential since chronic pathology over time can produce these sudden fatal outcomes without discernible patterns of symptoms beforehand. Understanding disease trajectory paths can help widen the intervention window and provide insights into warning signs, underlying disease etiology, and which lines of treatment are more promising in certain patients with predispositions or multimorbidities than others (Beaulieu-Jones et al., 2018; Oh et al., 2021).

Medical research recognizes common disease patterns where one condition often precedes another. Still, the sequence of events associated with an outcome is unknown for the vast majority of complex diseases (Hanauer & Ramakrishnan, 2012). An additional layer of complexity is introduced when we recognize the prevalence of multimorbidities, or the coexistence of two or more chronic diseases in an individual, in the progression of overall health. Most disease trajectory approaches fail to consider multiple lines of pathology that may be related and biologically interacting. This leads to building static disease trajectories based on the frequency of events rather than the co-occurrence of an outcome-precursor to an outcome of interest. To address these previous challenges and limitations, we propose a feature extraction algorithm **D**iseas**E T**rajectory f**E**ature extra**CT**ion (DETECT), that identifies individual-level trajectories for specific outcomes of interest. This preserves the meaningful relationships of conditions associated with the outcome by removing noisy variables while maintaining the relevant clinical features that define an individual patient’s disease trajectory.

## Methods

Jensen et al. have demonstrated that relative risk (RR) can be used to measure the strength of correlation between pairs of conditions in trajectory-building (Jensen et al., 2014). Relative risk is a measure of a certain event occurring in one group compared to the risk of the same event happening in another group. DETECT uses the relative risk framework to assess which events present in the data are predictive of an outcome. To build more accurate individual patient disease trajectories, selecting clinical events that signpost an outcome of interest would reduce noisy features and data dimensionality. A signpost code is a diagnosis that exhibits a RR > 1 and OR p-value > 0.05 and occurs in the disease trajectory in between the index diagnosis and the clinical outcome. DETECT implements a convergent RR based model to identify clinical features predictive of an outcome.

The input is 1) a matrix of individual-level, binary clinical variables with temporal information, 2) an index code or starting condition, and 3) a list of clinical outcomes of interest. **Fig. 1** depicts a workflow of the approach. The input contains only the first occurrence date associated with each corresponding health condition for each person; thus, every individual has a single date for each unique clinical variable in their chart. DETECT evaluates whether the presence of an intermediate condition is predictive of an outcome. If the intermediate condition meets the specified relative risk threshold RR > 1 and a p-value threshold of 0.05, it is considered a “signpost” code for that outcome. **Fig. 2** provides the convergent relative risk model used. Signpost codes are extracted from an individual’s raw health variables (diagnosis codes) string to produce a disease trajectory, which can be used in further predictive modeling or patient clustering analyses. To evaluate the validity of our approach, we simulated binary EHR data variables of varying degrees of individual-level noise. We benchmark DETECT by generating simulations of varying complexity and conducting sensitivity analyses.

**Fig. 1:**
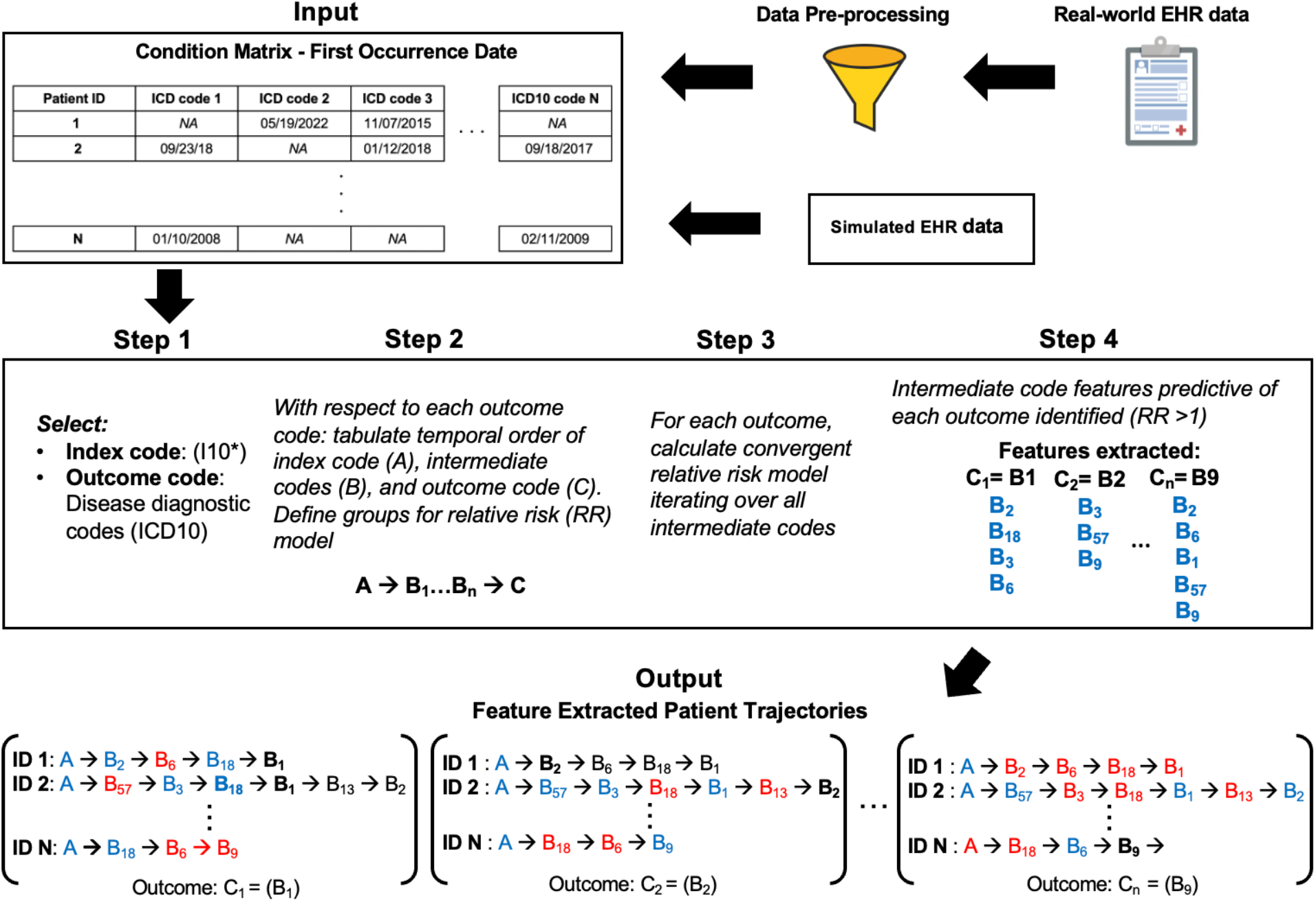
DETECT workflow. Input data is a matrix of first occurrence dates for each binary health variable for each individual. Real-world data can be pre-processed using a dimensionality reduction algorithm, or simulated data can be used. In step 1, we selected hypertension as our index code and outcome conditions (Fig. 6). Step 2 iteratively tabulates the temporal order of A (index code), B_n_ (intermediate codes), C (outcome) for each outcome. Step 3 calculates a convergent relative risk model for each outcome, testing the predictiveness of B with respect to C in people with and without B. Step 4 produces a list of predictive features (signpost codes) for each C. The output contains individual-level trajectories for each C with selected signpost codes (blue) from step 4, dropped codes (red), C (bold black), and the codes after the outcome are not returned (black). Pairwise time deltas (TΔ) are also provided.

**Fig. 2:**
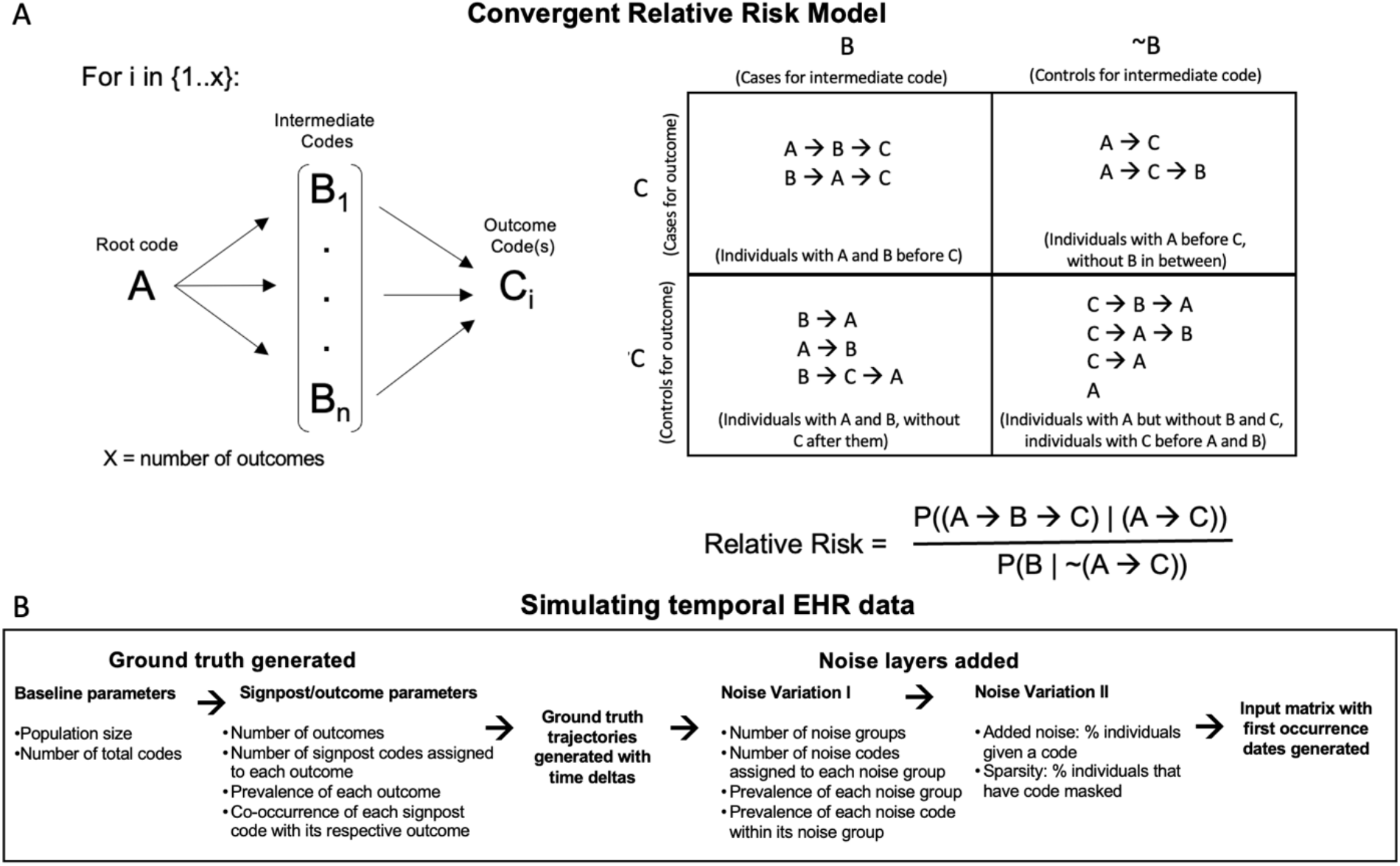
Convergent relative risk model and simulation generation. **A**. For each outcome (C), a relative risk calculation was done evaluating each intermediate code (B) to determine if it was predictive of C given a comparison between individuals who had both B and C and those who only had C. All individuals assessed as cases and controls for B and C had A as a prerequisite. Temporal placement of A, B, and C determined which quadrant an individual fell into. The different temporal configurations of A, B, and C in the EHR and how they are categorized are shown. In the relative risk equation, “A->B->C” includes both models in quadrant 1. **B**. The simulation generation pipeline in DETECT is outlined.

### Real-world EHR dataset pre-processing

We retrospectively obtained de-identified health records for 145,575 patients, extracting data from all patient encounters from 5 hospitals and their affiliated outpatient clinics in the Penn Medicine Health System between 1998 and 2022. We included patients who had at least 2 instances of hypertension (HTN) (I10*) diagnosis as a primary reason for the visit. Many severe outcomes are associated with HTN, yet we cannot predict which future disease states are likely in an individual diagnosed with HTN. The asterisk in I10* signifies that whichever code in the I10 hierarchy came first was used for the date of the first occurrence. We used the ICD-10 billing code system to represent health diagnoses. Any ICD-9 codes in patient charts were converted to ICD-10 using general equivalence mapping (Rhonda Butler, 2007). We excluded individuals for whom any I10* diagnosis was their first primary or secondary reason for visit in their chart to eliminate temporal bias introduced by patients who may have come to Penn as a specialty only after their HTN diagnosis elsewhere. Next, individuals with any codes for cardiac or respiratory congenital conditions were excluded to control for pathologies unrelated to hypertension. For the remaining cohort, any codes for external injury/harm and pregnancy-related conditions were also excluded to control for blood pressure related changes brought on by pregnancy or external factors. Codes were formatted to their 4-digit form based on the ICD hierarchy. To keep a primary diagnosis code at the individual level: 1) rare codes with a prevalence of ≤ 1% were only required to be present at least once, 2) common codes with >1% prevalence were required two or more occurrences in an individual to be kept (referred to as rule-of-two) (Hebbring, 2014). The secondary reason for visits (also known as “problem list” codes) were not evaluated for the cohort other than I10* as a criterion. The first occurrence of each unique ICD code for each individual was retained in the input data matrix where columns are ICD codes and rows are unique individuals (**Fig. 1**). 1385 4-digit ICD10 codes were included in this study.

### Trajectory model development

The DETECT user specifies a list of outcome diagnosis codes and the index code. The model is generated based on the classification of individuals into four quadrants of a contingency table for relative risk calculation (as shown in **Fig. 2**). All available codes that are not index or outcome are tested as intermediate codes. A separate model is generated for each clinical outcome; in other words, different features may be associated with or predictive of different outcomes within the same individual. Thus, the same individual could have different features extracted from their raw data to build a different trajectory for each outcome. The input matrix is used to categorize individuals into different quadrants of a contingency table (unique to each outcome): quadrant 1) individuals with the outcome code and the intermediate code, 2) individuals without the intermediate code and with the outcome code, 3) individuals without the intermediate code and the outcome code, and 4) individuals with the intermediate code and without the outcome code. This schema becomes more complicated when accounting for 1) irregularities and limitations of the EHR and 2) biological complexity, such as when an individual deviates from the simplistic A -> B -> C ordering. In our study, we did not require quadrant 1 individuals to have intermediate codes occur after index code HTN, only that HTN and the intermediate code occur before the outcome (**Fig. 2A**). This is because there were many individuals who had a HTN diagnosis after their intermediate code, and cases where the HTN and intermediate code were given on the same date. A time parameter allows the user to specify how many years max are allowed between an intermediate code and an outcome for the intermediate code to be evaluated in the relative risk model.

### Simulating EHR data ground truth

Because there are many imperceptible noise patterns in real-world EHR data, we benchmarked DETECT by generating simulated EHR data, and varying data structures by adding multiple levels of noise based on real-world data distributions. As part of DETECT, we have provided sensitivity-testing code to allow users to simulate ground truth temporal EHR data and introduce complexity through noise parameters (**Fig. 2B**). We evaluated how well the method could identify signpost codes despite added noise. Following is our methodology for generating ground truth with varying parameters; the subsequent section details our noise parameters. We first set the number of signpost codes a given outcome could have to between 50-300, the broad range accounting for clinical variation in complex traits. We then generated a data matrix with *N* rows and *M* columns where *N* is the population size or number of people with the index code, we tested population size between 50,000 and 150,000 (**Fig. 3D**). *M* is the number of codes we tested between 500 and 1,500 (**Fig. 4A**). We simulated between 10 and 60 outcomes (**Fig. 3C**). We sampled each outcome’s prevalence *p*_*outcome*_ from a uniform distribution between 0% and 10% prevalence (**Fig. 3A**). We assigned people to an outcome according to the chosen prevalence. Second, we sampled the number of signpost codes belonging to an outcome from a uniform distribution between 50 and 300 and assigned each signpost code a co-occurrence probability sampled from an exponential distribution with mean = 0.04 (**Fig. 3B**). Third, we determined the probability a signpost code would co-occur with its respective outcome code based on binomial variables generated representing each signpost code. Fourth, all instances of people being assigned codes were filled in with random time values preceding the time value at which the onset of the outcome occurred. Once this was completed for all outcomes, the ground truth trajectories were generated, with varying lengths for all individuals. Some individuals may have one, several, or no outcome codes; people with no outcomes also lack signpost codes up until this point.

**Fig. 3:**
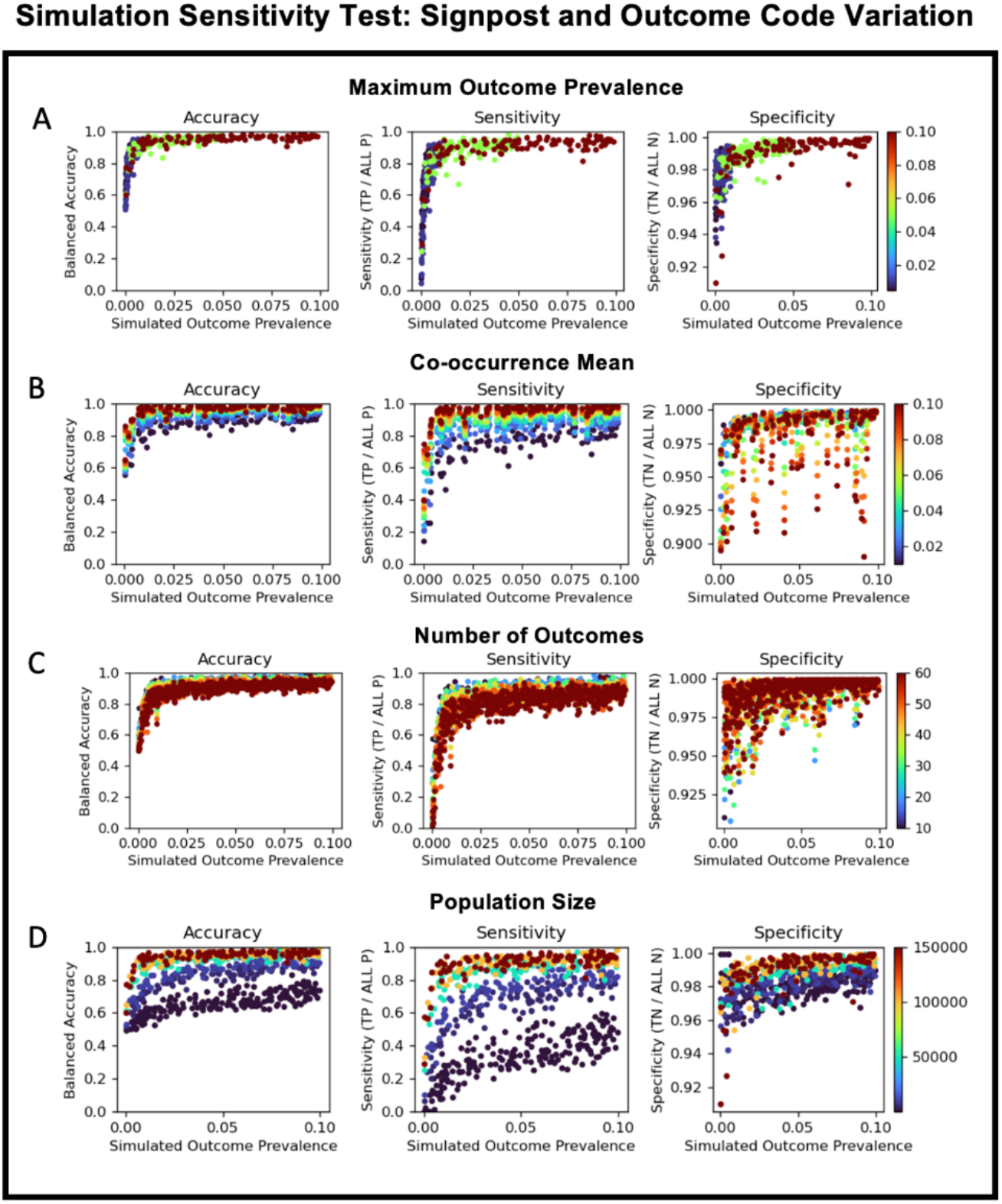
Sensitivity testing for outcome and signpost code variation. Variation introduced by changing **A**. outcome prevalence. **B**. co-occurrence of signpost code and outcome. **C**. number of outcomes, and **D**. population size are benchmarked through sensitivity, specificity, and accuracy calculation.

**Fig. 4:**
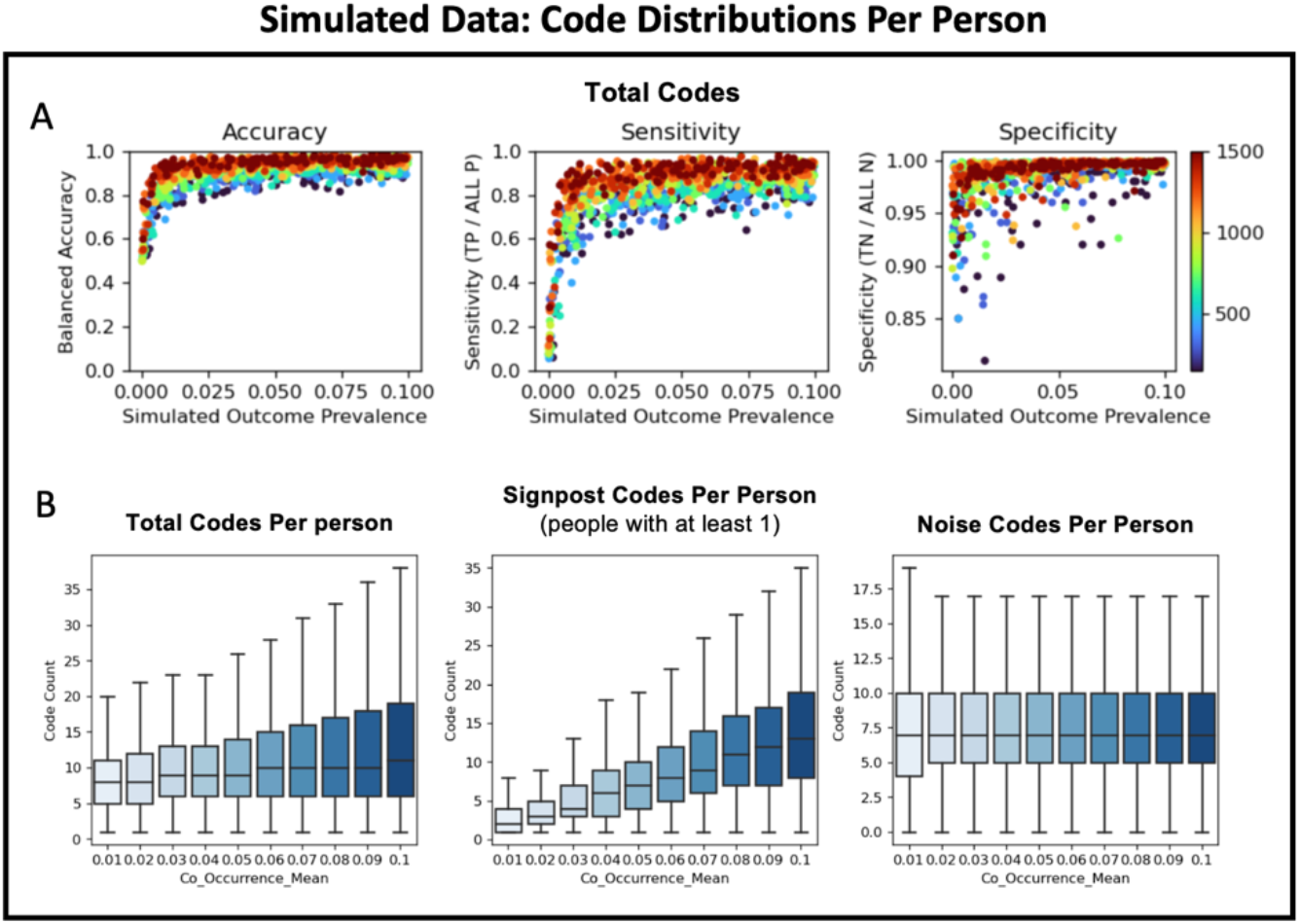
Simulated data code distributions. **A**. evaluates sensitivity, specificity, and accuracy as total number of codes increases, while **B**. shows the breakdown of total codes, signpost codes, and noise codes as a function of the co-occurrence of signpost codes with the outcome in individuals who have both.

### Simulating EHR data noise layers

Once we simulated the ground truth, we implemented two layers of noise variation to introduce realistic complexity into the data. Noise layer 1 consisted of simulated groups of codes as noise, representing unordered patterns of codes that may be due to aging, infections, medication side effects, etc. We simulated between 0-100 noise groups (**Fig. 5C**) in the data. Similar to the outcomes above, each noise group occurred in *p*_*noise*_ fraction of the population, with *p*_*noise*_ being sampled from a uniform distribution of 0% to 50%. The number of codes that would comprise a noise group was sampled from a uniform distribution between 5 to 300. All noise group codes were assigned a probability of occurring in a person belonging to a given noise group, sampled from an exponential distribution with mean = 0.01 (**Fig. 5B**). We applied the same concept of sampling binomial variables and assigning time values as in the outcome/signpost code assignments mentioned previously. The second layer of noise generated through parameter ‘Maximum Random Noise Fraction’ (**Fig. 5A**) was added to mimic both sparsity, which can be thought of as false negatives, and outcome-agnostic noise, or false positive appearances of codes. Codes are randomly assigned to people or are taken away from people. This can include signpost, outcome, or noise codes. The number of people for whom a code is added or masked depends on the number of people already assigned the code in the ground truth and first layer of noise, as well as a proportion sampled from a uniform distribution between 0% and 5%. For example, if 1000 people were assigned ICD code 1, 0-5% of those people would have code ICD code 1 removed from their data, and 0-50 new people would have code 100 inserted into their data at a random time point. The second layer of noise is not applied to the index code.

**Fig. 5:**
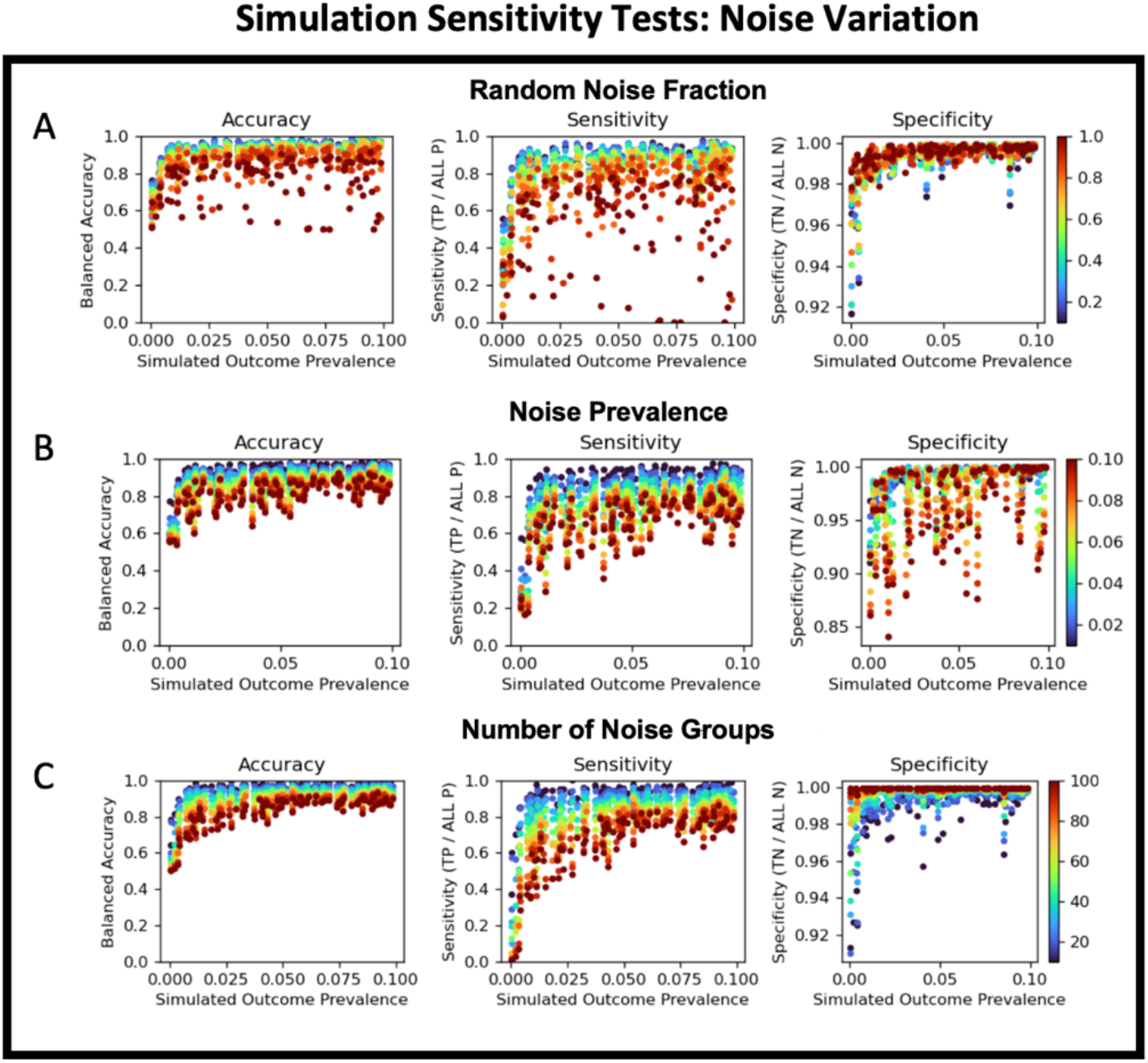
Sensitivity test on simulations with noise variation. Noise variation was introduce to increase complexity of simulated HER data. Accuracy, sensitivity, and specificity were calculated for parameters **A**. random noise scale. **B**. co-occurrence noise mean, and **C**. number of noise groups.

### Sensitivity testing in simulated EHR data

The output from DETECT is a table of all pairwise tests of relative risk between intermediate codes and outcomes, with co-occurrence counts, relative risk scores, and confidence information returned. A filtered list of intermediate codes with relative risk significantly greater than 1 (odds ratio p-value ≤ 0.05) are returned in a list for each outcome specified; these are signpost codes. The identified signpost codes for each outcome are tested against the known signpost codes that were embedded in the trajectories of that outcome. Comparing these two lists, we calculated the balanced accuracy ((sensitivity+specificity)/2), sensitivity (TP/(TP+FN)), and specificity (TN/(TN+FP)) to measure the correctness of feature selection. We benchmarked our approach additionally by parameterization. We tested a range of values for fixed and sampled variables, varying the noise level of the data to determine if the method was sensitive to a particular attribute.

### DETECT simulation parameters for benchmarking

**Table 1** contains parameters that the user can set to generate simulated temporal EHR data (binary variables) in the simulation part of the DETECT algorithm. To generate 879 simulation datasets for this study, approximately 200 single-threaded jobs were run concurrently, using up to 300 core hours and completed in 1.5hrs.

**Table 1:** User-defined parameters for generating simulated individual-level temporal EHR data in DETECT. The max of a uniform distribution is denoted as *b*. The mean of an exponential distribution is denoted as *1/λ*.

| Category | Parameter Name | Parameter Description | Default |
| --- | --- | --- | --- |
| Outcome | Number of Outcomes | Number of outcomes tested | 10 |
| | Maximum Outcome Prevalence | $b$ of uni. dist. for sampling prevalence | 0.1 |
| | Co-Occurrence Mean | $1/\lambda$ of exp. dist. for sampling code probabilities | 0.04 |
| Noise Layer 1 | Number of Noise Groups | Number of noise groups simulated | 20 |
| | Maximum Noise Group Prevalence | $b$ of uni. dist. for sampling prevalence | 0.01 |
| | Noise Prevalence Mean | $1/\lambda$ of exp. dist. for sampling code probabilities | 0.5 |
| Noise Layer 2 | Maximum Random Noise Fraction | $b$ of uni. dist. for sampling noise to add/subtract | 0.05 |
| Baseline | Population Size | Number of individuals in the population | 150,000 |
|  | Number of Codes | Number of codes simulated | 1500 |

### Additional Output from Application of the method to real-world EHR Data

A file with trajectories for all individuals is produced separately for each outcome, containing the extracted signpost features, index code, and outcomes. A JSON file with time deltas between each feature is also produced to accompany each trajectory file. Code for the analysis is available: https://github.com/5inghalp/DETECT.

## Results

In this study, we demonstrate the ability and utility of our algorithm DETECT to generate patient-level trajectories using simulated and real-world EHR data. Our real-world analysis uses EHR data from the Penn Medicine health system on 145,575 patients diagnosed with HTN. HTN was defined as our index code and we generated and evaluated trajectories for eight outcomes of interest (**Fig. 6**).

**Fig. 6:**
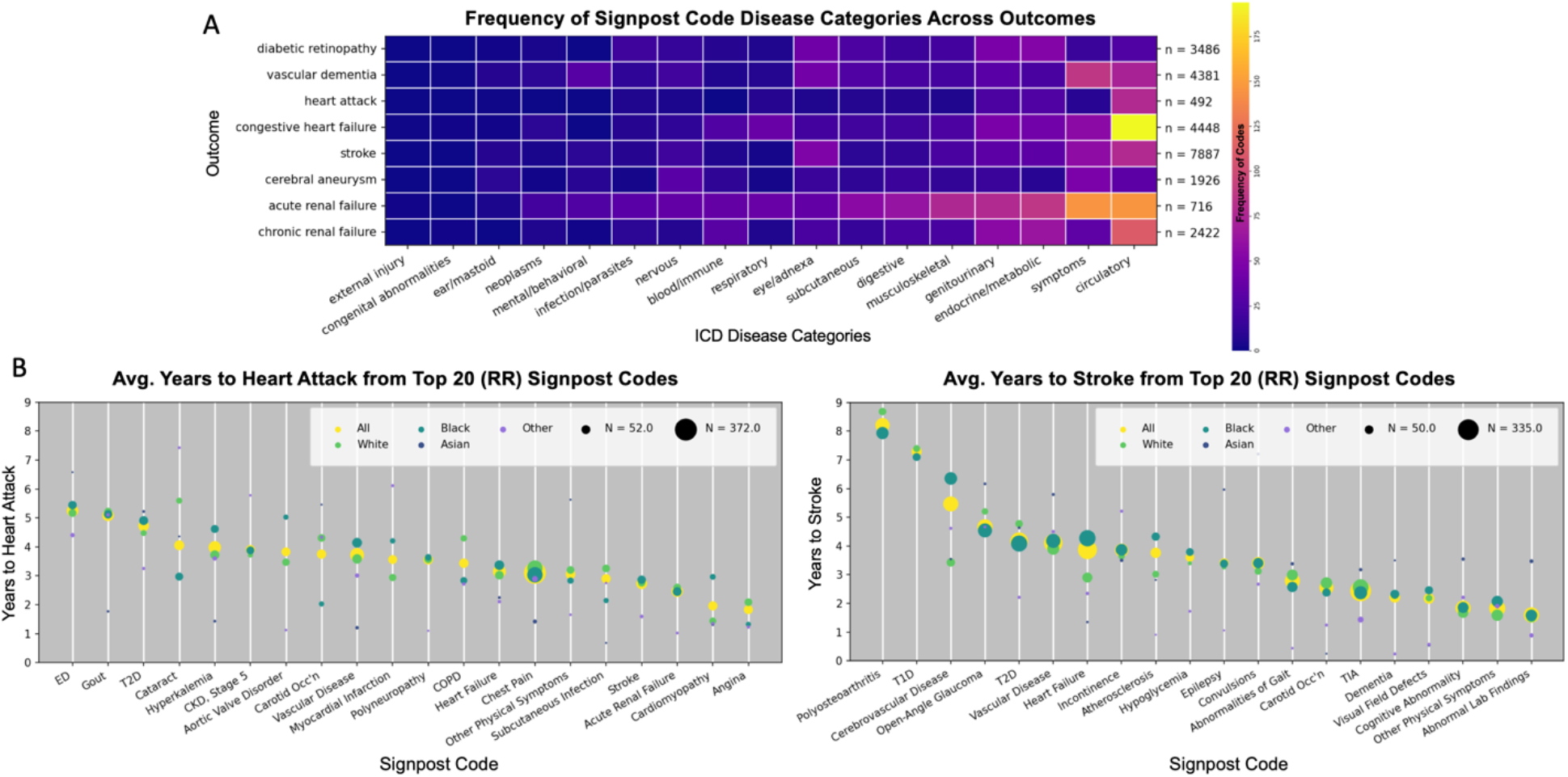
Frequency of signpost code categories and time to outcome for select outcomes. **A**. The heatmap indicates the frequency of signpost ICD codes (grouped to ICD disease categories based on the latest international version) for each outcome of interest. Sample sizes are indicated for each outcome. **B**. Trajectory plot shows average time (in years) to outcome for the top 20 highest RR signpost codes detected for heart attack (left) and stroke (right). In each plot, the color represents self-reported ancestry and the size of the circle corresponds to sample size.

### Simulations and sensitivity testing

We benchmarked our approach by conducting simulation studies in which the ground truth relationship between signpost variables and outcomes were embedded. Sensitivity, specificity, and accuracy were calculated based on whether DETECT could correctly identify true signpost-outcome relationships. **Fig. 3** shows sensitivity analyses benchmarking the ability of DETECT to identify signpost codes when complexity is introduced into the ground truth. First, we evaluated the parameters affecting the dimensionality and quality of ground truth in the simulated EHR data. DETECT produces higher accuracy when max prevalence of outcome codes is greater than 1% (**Fig. 3A**). Rare events occur in few people, therefore there are few signpost codes associated with <1% conditions, and the chance of selecting a non-predictive (noise or neutral) code is higher. In **Fig. 3B**, accuracy and sensitivity follow a positive increasing trend as co-occurrence of signpost codes and outcomes increases. Specificity, however, shows an upward “waterfall effect”, suggesting an increase in selection of false positive signpost codes. This is because reduced co-occurrence with the outcome code may cause increased occurrence with neutral or noise codes with the outcome, leading to an increased RR for false relationships. In **Fig. 3D**, as the number of outcome codes in the dataset increases, the accuracy and sensitivity decreases, this is expected as increased outcomes result in increase in noise codes, as well as signpost codes. Because a single signpost code can be linked to multiple outcomes, a high level of crossover can reduce the ability to correctly identify true signpost-outcome relationships. The specificity, however, increases as the number of outcomes increases. This is likely because codes in a trajectory have a greater chance of being a signpost code for an outcome if there are multiple outcomes, reducing false positive rates. In the simulated data, accuracy, sensitivity, and specificity all increase as the fixed number of codes increases (**Fig. 4A**). A distribution signpost and noise code counts is shown in **Fig. 4B** as co-occurrence of signpost code and outcome is increased. Similarly, when the population size exceeds 50,000 people, accuracy exceeds 80% (**Fig. 3D**). This is most likely due to a higher degree of co-occurrence increasing the total frequency of the signpost codes. This leads to an increase in both signpost codes per person and overall codes. More codes would increase the chances of selecting nonsignpost or noise codes, resulting in a greater false positive rate.

We evaluated DETECT performance after addition of noise parameters, introducing realistic complexity into the simulated EHR data. Increasing both the level of noise layer 2, which adds or masks codes to mimic noise and sparsity, and increasing number of noise groups, results in a decreased accuracy and sensitivity (**Fig. 5A** and **Fig. 5C**). This is due to the influx of nonsignpost-outcome instances in individuals with the outcome, resulting in higher RR values than actual signpost-outcome occurrences. This causes a higher false negative rate. Interestingly, the specificity increases as both noise layer 2 and number of noise groups increase. This is likely because when there are fewer noise groups, and therefore fewer noise codes, the frequency of the noise code may not be high in individuals with the outcome. This would cause RR to still be high for some of the noise codes, thus being selected as signpost codes for outcomes, when they are not. As prevalence of noise codes within noise groups increases, a higher false positive rate emerges, as is expected (**Fig. 5B**).

### Real-world data trajectory analysis

We applied DETECT on a real-world dataset from Penn Medicine EHR, on a cohort of 145,575 individuals with hypertension diagnosis codes. We generated trajectories for the following outcomes of interest defined by ICD-10 codes: diabetic retinopathy (E11.3), vascular dementia (F01*), myocardial infarction (I21*), congestive heart failure (I50*), cerebral infarction (I63*), cerebral aneurysm (I67.1), acute kidney disease (N17*), and chronic kidney disease (N18.4). The heatmap in **Fig. 6A** shows the frequency of the extracted signpost codes, mapped to ICD10 disease categories, for each outcome of interest. We selected outcomes that are known severe comorbidities of HTN. Expected relationships are identified in this analysis as positive controls. For example, a high frequency of circulatory signpost codes are predictive of congestive heart failure. A medium frequency of eye/adnexa signpost codes are predictive of diabetic retinopathy, stroke, and vascular dementia, all shown in the literature to be associated with ocular symptoms (Ho et al., 2009; Hwang et al., 2021; Wong et al., 2020). An application for how individual-level DETECT trajectories can be leveraged to ask temporal questions about order or quality of disease precursors at a population level is shown in Fig. **6B**. For outcomes of heart attack and stroke, the average time delta (years) between the top 20 significant relative risk signpost codes and the outcome is shown. This analysis is stratified by self-reported ethnicity. The pattern of symptoms emerging over time is relative to the outcome, irrespective of personal timelines. In addition to expected aging related codes, both outcomes had phenotypes that would be considered “positive controls”. COPD, carotid occlusion, aortic valve disorder, and chest pain, among others, are all considered known comorbidities for heart attack. Glaucoma, transient ischemic attack, cerebrovascular disease, and atherosclerosis are known symptoms or comorbidities of stroke. Median age for stroke individuals was 71.6 years and median age for heart attack individuals was 69.9 in our dataset.

## Discussion

Ground truth relationships between most clinical events or variables (including but not limited to labs, medications, vitals, ICD codes, CPT codes, demographics, etc.) are not known in the context of real patient data, posing a challenge for methods modeling with clinical information. This is in part due to artifacts of the EHR such as establishment, clinician, and insurance-specific coding practices that do not reflect true pathology. Absence of patient visits cannot be interpreted as periods of high health, however, more often than not that is the interpretation made. Medical knowledge can explain biological associations in well-studied pathologies, but it does not prescribe a replicable pattern or sequence of events observable across patients for complex diseases. DETECT uses a data-driven approach to extract clinical binary features that are predictive of outcomes of interest. Resulting patient trajectories can more intuitively be integrated with other recurring clinical data types such as labs, medications, and CPT codes. Our sensitivity analyses benchmarking DETECT with simulated data of varying complexity show high accuracies on average above 80% for predicting meaningful signpost codes. Simulated individual-level data contained trajectories of varying number of outcomes, noise codes, true signal/signpost codes, time deltas, and sparsity. Our comprehensive approach of generating realistic binary EHR diagnostic codes comprises the first part of our algorithm.

We applied DETECT to real-world data in order to generate patient trajectories for eight severe outcomes. **Fig. 6A** shows the frequency distribution of signpost codes (mapped to ICD disease categories) for each outcome of interest. Interestingly, there is a difference in patterning between acute and chronic renal failure, with acute renal failure having many more diverse predictors across the disease categories. One reason for this could be that there are more prominent disease precursors in different pathologies that can lead to an acute renal failure state, whereas chronic renal conditions are slow moving over time. There are some expected results such as vascular dementia having codes across mental/behavioral, eye/adnexa, and circulatory categories, all affecting the head. Diabetic retinopathy exhibited a similar pattern, but also included genitourinary and metabolic codes. Further work can be done examining patterns within disease categories for additional granularity in symptoms over time.

To demonstrate how trajectories from DETECT can be leveraged to evaluate temporal trends in EHR data, we ordered the top 20 signpost codes (with minimum 50 cases) by average time delta for two outcomes, heart attack and stroke, based on RR value (**Fig. 6B)**. Over the span of 9 years prior to the outcomes, average placement of detected signpost predictor codes is shown. Individuals with epilepsy have been shown to have stroke in a higher proportion than individuals without epilepsy, however epilepsy has not been identified as a predictor of stroke (Chang et al., 2014). Our temporal analysis shows that not only epilepsy, but also convulsions, are predictors of stroke. As are ocular phenotypes including open-angle glaucoma and visual field defects, both of which can occur due to high blood pressure.

The major strength of DETECT is to analyze longitudinal EHR data in a high-throughput approach to derive temporal trajectories for disease of interest. However, we acknowledge there are following limitations to the current methodology. First, our approach to use “date of the first occurrence” as “condition onset,” works well to represent ICD codes as binary variables because they represent the diagnosis of a certain condition at a given time. However, a similar approach to represent recurring variables such as medication, CPT procedure codes, and vitals can be challenging, as without contextual information it can lead to underlying bias in the disease trajectories. Second, given the larger sample size of our study, we used p-value < 0.05 and RR > 1 to select temporality for pairs of diseases. However, with smaller cohorts, a p-value threshold may be too restrictive to detect important signals. Power calculations and additional simulations are needed to determine this. Further work is needed to incorporate non-binary and recurring EHR data variables. Future directions for this work would include developing a predictive modeling component to DETECT to conduct clinical event prediction in disease trajectory sequences. Specifically, leveraging time deltas between conditions at a population scale can produce insights about time estimations for the next clinical event for a patient. Understanding the next likely disease an individual is at risk for can widen the clinical intervention window available in lines of treatment.

## Data Availability

All data produced in the present work are contained in the manuscript.

https://github.com/5inghalp/DETECT

## Acknowledgements

MDR is supported by R01HL141232, R01GM138597, and UL1-TR-001878. AV and DK are supported by NIGMS R01-GM138597. PS is supported by F31 AG069441-01. This manuscript was submitted for consideration for the AMIA 2023 Informatics Summit and the decision is pending.

## Notes

### Competing Interest Statement

The authors have declared no competing interest.

### Funding Statement

MDR is supported by R01HL141232, R01GM138597, and UL1-TR-001878. AV and DK are supported by NIGMS R01-GM138597. PS is is supported by F31 AG069441-01

### Author Declarations

Institutional Review Board of the University of Pennsylvania gave ethical approval for this work through IRB Protocol #829861. The project qualified as exempt.

